# Childhood Mental Health and Body Mass Index as Mediators of Genetic Risk for Eating Disorders

**DOI:** 10.64898/2026.03.13.26347917

**Authors:** Chaoyu Liu, Jiayi Xu, Adrianna Kępińska, Yen-Feng Lin, Eating Disorders Working Group of the Psychiatric Genomics Consortium, Gerome Breen, Jonathan R.I. Coleman, Cynthia M. Bulik, Laura M. Huckins

**Affiliations:** Division of Molecular Psychiatry, Department of Psychiatry, Yale University School of Medicine, New Haven, USA; Social, Genetic and Developmental Psychiatry Centre, Institute of Psychiatry, Psychology & Neuroscience, King’s College London, London, UK; The Charles Bronfman Institute for Personalized Medicine and Department of Genetics and Genomic Sciences, Icahn School of Medicine, New York, USA; Department of Genetic and Genomic Sciences, Icahn School of Medicine, New York, USA; Center for Neuropsychiatric Research, National Health Research Institutes, Miaoli, Taiwan; National Institute for Health and Care Research Maudsley Biomedical Research Centre, South London and Maudsley National Health Service Trust, London, UK; Department of Psychiatry, University of North Carolina at Chapel Hill, Chapel Hill, USA; Department of Medical Epidemiology and Biostatistics, Karolinska Institute, Stockholm, Sweden; Department of Nutrition, University of North Carolina at Chapel Hill, Chapel Hill, USA

## Abstract

**Importance:** Eating disorders (EDs) are heritable, yet the developmental pathways through which genetic liability manifests in early life remain unclear.

**Objective:** To investigate the associations between genetic liability for anorexia nervosa (AN) and binge eating (BE) and disordered eating behaviors (DEB) across childhood, and to identify the mediating roles of metabolic and psychosocial traits.

**Design, Setting, and Participants:** This longitudinal observational study used genomic and behavioral data from the Adolescent Brain Cognitive Development ^SM^ (ABCD^®^) Study, a multisite, population-based cohort of children recruited between 2016 and 2018 at ages 9 to 10 years from 21 research centers across the United States. A three-wave temporal design was employed, utilizing data from baseline (T0), Year 1 (T1), and Year 2 (T2) follow-ups. Primary analyses focused on 5,618 participants of genetically inferred European (EUR) ancestry, with exploratory analyses conducted in a diverse sample of 9,132 participants.

**Exposures:** Polygenic scores (PGS) for AN and BE were calculated using summary statistics from the most recent genome-wide association studies. Mediators included BMI, ADHD, anxiety/depression, and social problems from the Child Behavioral Checklist assessed at Year 1 follow-up (T1).

**Main Outcomes and Measures:** Parent reported DEB symptoms via the Kiddie Schedule for Affective Disorders and Schizophrenia (KSADS). For longitudinal association analyses, DEB were pooled across T0, T1 and T2 to assess the relationship between genetic liability and childhood symptom severity. For mediation analyses, DEB at T2 follow-up were used to ensure a clear temporal sequence between mediators at T1 and the outcomes.

**Results:** Among 5,618 EUR participants (mean [SD] age, 9.91 [0.62] years; 47% female), longitudinal association models revealed that higher AN-PGS was associated with increased AN symptoms, while BE-PGS was associated with increased BE and AN symptoms. These patterns were largely consistent in exploratory cross-ancestry analyses. Mediation analyses showed that BMI mediated genetic risks across sexes, while ADHD and anxiety/depression symptoms emerged as additional mediators in females.

**Conclusions and Relevance:** Genetic liabilities to AN and BE contribute to childhood DEB through sex-dependent pathways, highlighting the developmental continuity of ED risk from childhood. Integrating genetic profiles with behavioral markers may facilitate early identification and support multifaceted interventions.

**Key points Question:** Do genetic risks for anorexia nervosa (AN) and binge eating (BE) contribute to childhood disordered eating behaviors, and what mechanisms mediate these effects?

**Findings:** In this longitudinal study of 5,618 children of European ancestry, AN polygenic scores (AN-PGS) were associated with early AN symptoms, while BE-PGS showed transdiagnostic associations with both AN and BE symptoms. These links were mediated by BMI and psychosocial traits, including sex-specific pathways through ADHD and anxiety/depression symptoms in females.

**Meaning:** Our findings suggest that genetic liability to eating disorders manifests early in life through distinct metabolic and psychosocial pathways, highlighting a window for sex-specific targeted prevention.

## Introduction

Disordered eating behaviors (DEB) in childhood often persist or worsen during adolescence and early adulthood, and are therefore considered strong predictors of later eating disorders (EDs) ^1^. However, it remains unclear whether childhood DEBs and adult EDs share common genetic risks, or whether additional factors likely contribute across different developmental stages ^2^. Understanding these pathways is critical for early identification and intervention.

Genome-wide association studies (GWAS) have deepened our understanding of the genomic architecture of EDs. Major collaborative efforts by the Eating Disorders Working Group of the Psychiatric Genomics Consortium (PGC-ED) have identified genome-wide significant loci associated with anorexia nervosa (AN) ^3^ ^4^ ^5^ and binge eating (BE) ^5^. In addition, these studies have highlighted the developmental nature of EDs, revealing shared genetic liability with pubertal timing and age at menarche, and implicating biological processes relevant to growth and maturation ^4,6,7^. However, most genetic research to date has focused on adults, leaving the pathways linking genetic liability, childhood DEB and their continuity to adult EDs poorly explored.

Polygenic scores (PGS), which aggregate the effects of numerous common genetic variants identified in GWAS, are increasingly applied to index individual genetic liability for complex traits ^8^. While AN-PGS have been associated with EDs ^9^, and with ED-related traits ^10^ in adults, the evidence in childhood remains scarce. To date, studies examining AN-PGS in youth cohorts have yielded inconsistent or null associations with DEB ^6^ ^11^ ^12,13^ ^14^, while such findings have not yet emerged for BE-PGS. The mixed findings for AN-PGS may reflect limited statistical power of earlier GWAS and the developmental heterogeneity and dynamic nature of childhood DEB. For example, early-life overeating has been linked to both BE- and AN-related symptoms in young adulthood ^15^, highlighting the complex, sometimes transdiagnostic trajectories of childhood DEB. In addition, phenotypic and genetic studies implicate traits such as BMI, attention deficit/hyperactivity disorder (ADHD), and depression in the emergence of childhood DEB ^16^, suggesting that metabolic and psychosocial pathways jointly shape risk. Overall, these findings highlight the importance of studying not only direct genetic effects but also the interplay between genetic liability and intermediary traits for childhood DEB.

Sex differences are a critical aspect of ED risk. While clinical features often show similarities, notable differences between sexes exist in epidemiological findings such as prevalence and developmental trajectories, and underlying neural and biological risk factors ^17,18^. These divergent pathways are particularly evident in the influence of gonadal hormones during sensitive developmental windows, such as the pubertal transition, which may modulate the expression of genetic liability and influence the emergence of sex-specific phenotypes ^19^. These disparities extend to specific symptoms and response to sociocultural pressure ^20^. For example, males are more likely to engage in excessive exercise, muscle-building behaviors or binge eating, often influenced by weight-related teasing or societal ideas of muscularity ^21,22^, whereas females more commonly exhibit restrictive eating and compensatory behaviors driven by body dissatisfaction and pressure to be thin ^23^ ^24^. Comorbidities such as depression and anxiety, which are more prevalent in females, further exacerbate eating pathology and complicate disease outcomes ^16,25^. Biological Family-based research also suggests sex-specific heritability patterns.

Twin studies have shown higher heritability estimates in females, and an earlier activation of genetic influences in males, particularly around pre-puberty ^19,26^. Molecular genetic findings further mirror this: for example AN-PGS predicted AN symptoms only in adolescent females ^12^. Together, these multidimensional differences shaped by behavioral, psychosocial and biological mechanism explain different susceptibility to EDs by sex and emphasize the necessity of sex-stratified frameworks in developmental and genetic studies of EDs.

This study examined how genetic liabilities for AN and BE (hereafter referred to as EDs) predict childhood DEB using longitudinal data from the Adolescent Brain Cognitive Development (ABCD) Study and the latest, well-powered GWAS of AN and the first GWAS of BE. We investigated whether associations between ED-PGS and childhood DEB were mediated by anthropometric and childhood mental health conditions, and whether these mediation pathways differed by sex. We hypothesize that the updated ED-PGS would demonstrate stronger predictive utility and that sex-specific mediation would reveal distinct developmental mechanisms for ED risk. Primary analyses focused on individuals of European (EUR) ancestry to align with the discovery GWAS, with exploratory analyses in non-EUR participants to evaluate cross-ancestry consistency.

## Methods

### Study Participants

We used data from the ABCD study 5.1 release, a prospective longitudinal study of 11,878 children recruited from 21 sites across the United States around 9-10 years of age. The ABCD cohort was designed to approximate the sociodemographic characteristics and population structure of the United States ^27^. Written informed consent was obtained from parents/primary caregivers. Details about the cohort profile, data collection and genotyping protocol of the ABCD study are available elsewhere ^27,28^. This study followed the Strengthening the Reporting of Observational Studies in Epidemiology (STROBE) reporting guidelines.

### Phenotypic Measures

#### Disordered eating behavior (DEB) Symptoms

DEB symptoms were derived from the parent-report Kiddie Schedule for Affective Disorders and Schizophrenia (KSADS) ^29^ ED module. AN symptoms were assessed using 3 Likert-scale items (0: not at all to 4: nearly every day): “preoccupation with gaining weight”, “control weight by throwing up”, and “control weight by other methods (exercise, taking pills, restrict food)”.

Item scores were summed to index AN severity. BE symptom level was calculated as the product of two items: BE frequency “how often the child binge eats/lost control of eating” (range 0 to 4) and associated discomfort “how much discomfort does binge eating cause” (range 0 to 10) to derive a composite severity score. Item-level missingness was <1% and addressed using mean substitution. DEB symptoms at three time points (Baseline [T0], Year 1 follow-up [T1], and Year 2 follow-up [T2]) were utilized in longitudinal association analyses using a multilevel framework to account for repeated measures. For the mediation analyses, only symptoms at T2 were used to establish a clear temporal sequence between the mediators and the outcome.

#### Mediators: BMI and psychosocial traits

We focused on a set of theoretically informed mediators: child BMI, ADHD, anxiety/depression and social problems, all of which have been previously associated with ED risk both phenotypically and genetically ^16,30^. We used the standardized summary scores of Anxious/Depressed, ADHD and Social Problems syndrome subscales of the Child Behavior Checklist (CBCL) at T1 as mediators ^31^. Participants’ BMI was calculated based on the reported height and weight at annual follow-up. Extreme BMI values (<10 or >40) were recoded as missing. BMI was then converted to z-scores based on the 2000 Centers for Disease Control (CDC) growth chart reference ^32^ using *childsds* R package ^33^. We performed two models to examine BMI mediation: first, we used standardized BMI at T1 as the mediator and in the second model, we tested the change in BMI (ΔBMI) between baseline and T2 while controlling for baseline BMI.

### Genetic data processing and quality control

This study used the imputed genotyped data of the ABCD 5.1 release. Details of genotyping and TOPMED imputation details can be found in previous publications ^28^. In brief, the imputation was performed using standard protocols provided by the TOPMED Imputation Server, based on TOPMED reference panel. The TOPMED reference panel was chosen due to its diverse ancestry representation, which closely aligns with the characteristics of the ABCD samples. Due to the diverse and admixed population structure of the ABCD samples, we first used ADMIXTURE software ^34^ and 1000 Genomes Project Phase 3 reference populations ^35^ to infer individual genetic ancestry. Among 11,666 samples with genotyping data, 7,240 were assigned as European (EUR), 2,111 as American (AMR), 1880 as African (AFR), 216 as East Asian (EAS) and 49 as South Asian (SAS). A total of 170 participants were assigned as admixed sample and were excluded from further analyses. After genetic ancestry assignment, QC procedures were performed in each ancestral groups using PLINK (v.1.9). Individuals with genotype missingness >2%, sex mismatch, or excess heterozygosity (> 3 SD) were excluded. SNPs were excluded if they had a missingness rate >5%, minor allele frequency <1%, or Hardy-Weinberg equilibrium p < 1×10 . In the cases of related individuals, one individual with higher data missingness in a related pair (IBD pi-hat>0.2) was dropped. A total of 9,132 unrelated participants passed quality control, including 5,619 EUR, 1,727 AMR, 1,551 AFR, 190 EAS and 45 SAS.

#### Genotype-derived principal components

We implemented two complementary principal component analyses (PCA) to derive 2 sets of genotype-based principal components (PCs) on LD independent variants (r^2^<0.1, 500 SNP windows) for use in separate downstream analyses. First, for analyses restricted to participants of EUR ancestry, we generated ancestry-specific PCs using PLINK (v.1.9) with reference to the 1000 Genome Project ^35^ to use as covariates in EUR-specific analyses controlling for within-population structure. Next, to model genetic structure in the full, cross-ancestry sample, we applied PC-AiR ^36^, which estimates population structure while accounting for cryptic relatedness using a kinship matrix generated with the KING-robust method ^37^. For downstream analyses involving the **full** ABCD cohort, we used PCs derived from PC-AiR but restricted analyses to genetically unrelated individuals to minimize confounding from sample relatedness.

#### Polygenic score (PGS) calculation

PGS were computed for AN (N= 24,223 cases, 1,243,971 controls) and BE (N= 39,279 cases and 1,227,436 controls) using summary statistics from the largest available GWAS ^5^, which included participants of EUR ancestry. Given the multi-ancestral composition of the ABCD cohort, PGS were generated using PRS-CS ^38^, a Bayesian polygenic model that accounts for linkage disequilibrium (LD) variation across ancestries. PGS were generated and standardized (z-scores) within each ancestry group.

Because some BE-GWAS cases overlapped with AN-GWAS cohort, sensitivity analyses were performed using PGS derived from a BE-GWAS subset excluding AN-ascertained cohorts (BE-NAN, N= 34,304 cases and 1,180,607 controls) to examine if associations were driven by AN-related effects.

### Statistical Analyses

#### Associations between ED-PGS and childhood DEB

All continuous variables were standardized (mean=0, SD=1) before analyses. All analyses were conducted in R (v4.4.1) ^39^ and were stratified by sex. We fit generalized linear mixed models with random intercept to account for within-subject correlation (R package *glmmTMB)* ^40^ to investigate the associations between ED-PGS and DEB measured across baseline, T1 and T2.

Given that the discovery GWAS for EDs only included participants of EUR ancestry, primary analyses were conducted in the EUR subgroup. ED-PGSs were residualized on sex, 20 ancestry-specific PCs and genotype plates prior to association analyses. We then extended analyses to the full ABCD sample. In these analyses, ED-PGS were residualized on sex, 35 PCs derived with PC-AiR, and genotyping plate. All association models were controlled for age and self-reported race/ethnicity as covariates. Confidence intervals were estimated via nonparametric bootstrapping with 1,000 resamples to ensure robustness of the estimates. To correct for multiple testing, false discovery rate (FDR) adjustment of p=0.05 was applied.

#### Mediation between ED-PGS and childhood DEB

Mediation analyses were conducted using the R package *mediation* ^41^. We were interested in the average causal mediation effect (ACME) as it reflects differences in the outcomes when the mediator assumes different values between exposure and control conditions (i.e. differences between the observed outcome and the counterfactual outcome)^42^. In these models, PGS served as the exposure, T1 phenotypic traits (BMI and CBCL subscales) served as mediators, and DEB symptoms at T2 served as the outcome to satisfy the temporal requirements for mediation inference. Potential confounders including age at T1 and self-reported race ethnicity (in the full sample) were controlled in the model. Standard errors and 95% confidence intervals were derived using non-parametric bootstrapping with 1,000 simulations to provide robust inference under possible non-normality of data distribution.

## Results

### Eating Disorder symptoms are present in ABCD

Descriptive statistics for participants of EUR ancestry (n=5,618) and the full sample (n=9,129) are in Table 1 and the distribution of AN and BE scores is shown in Supplementary Figure 1. Wilcoxon rank-sum tests showed significant sex differences consistent with epidemiological patterns ^17^: females exhibited significantly higher AN symptoms across the follow-up (p<0.001), whereas males displayed higher levels of ADHD symptoms (p<0.001).

**Table 1.** Descriptive demographics of the study sample.

|  |  | EUR subgroup |  | Full sample |  |
| --- | --- | --- | --- | --- | --- |
|  |  | Male | Female | Male | Female |
| measure | timepoint |  |  |  |  |
| N | baseline | 3,003 | 2,615 | 4,797 | 4,332 |
|  | T1 | 2,905 | 2,520 | 4,548 | 4,070 |
|  | T2 | 2,852 | 2,462 | 4,458 | 3,970 |
| Age (mean, sd) | baseline | 9.92 (0.63) | 9.90 (0.62) | 9.92 (0.62) | 9.89 (0.62) |
|  | T1 | 10.93 (0.64) | 10.91 (0.64) | 10.93 (0.63) | 10.90 (0.64) |
|  | T2 | 12.03 (0.67) | 12.01 (0.67) | 12.03 (0.66) | 12.01 (0.67) |
| BMI (mean, sd) | baseline | 17.85 (3.33) | 17.91 (3.51) | 18.58 (3.98) | 18.80 (4.20) |
|  | T1 | 18.74 (3.71) | 18.73 (3.79) | 19.47 (4.33) | 19.74 (4.62) |
|  | T2 | 19.54 (4.13) | **19.78 (4.12)\*\*** | 20.28 (4.58) | 20.75 (4.84) |
| AN symptom<br>(mean, sd) | baseline | 0.27 (0.68) | **0.34 (0.73)\*\*\*** | 0.32 (0.80) | **0.39 (0.84)\*\*\*** |
|  | T1 | 0.39 (0.79) | **0.44 (0.86)\*** | 0.44 (0.88) | **0.49 (0.92)\*\*\*** |
|  | T2 | 0.45 (0.91) | **0.50 (0.93)\*\*\*** | 0.46 (0.92) | **0.51 (0.97)\*\*\*** |
| BE symptom<br>(mean, sd) | baseline | 0.25 (1.56) | 0.24 (1.50) | 0.34 (1.93) | **0.29 (1.79)\*\*** |
|  | T1 | 0.33 (1.63) | 0.40 (1.95) | 0.43 (2.02) | 0.43 (2.11) |
|  | T2 | 0.33 (1.73) | 0.38 (2.04) | 0.38 (1.94) | 0.36 (2.00) |
|  |  | Male | Female | Male | Female |
| measure | timepoint |  |  |  |  |
| Anxiety/Depression symptoms (mean, sd) | baseline | 2.68 (3.16) | 2.74 (3.14) | 2.57 (3.09) | 2.59 (3.09) |
|  | T1 | 2.76 (3.21) | **2.90 (3.24)\*\*** | 2.57 (3.11) | 2.65 (3.12) |
|  | T2 | 2.39 (2.96) | **2.84 (3.29)\*\*\*** | 2.22 (2.90) | **2.50 (3.09)\*\*\*** |
| ADHD symptoms (mean, sd) | baseline | 3.51 (3.75) | **2.41 (3.05)\*\*\*** | 3.62 (3.80) | **2.47 (3.13)\*\*\*** |
|  | T1 | 3.43 (3.73) | **2.41 (3.08)\*\*\*** | 3.46 (3.73) | **2.42 (3.11)\*\*\*** |
|  | T2 | 3.31 (3.63) | **2.33 (3.02)\*\*\*** | 3.30 (3.60) | **2.26 (2.95)\*\*\*** |
| Social problems (mean, sd) | baseline | 1.63 (2.37) | **1.44 (2.09)\*** | 1.75 (2.41) | **1.55 (2.19)\*\*** |
|  | T1 | 1.53 (2.25) | 1.44 (2.11) | 1.58 (2.25) | 1.48 (2.16) |
|  | T2 | 1.36 (2.14) | 1.30 (2.01) | 1.38 (2.16) | 1.30 (2.00) |

1. Full sample comprises participants from European ancestry (n=5618), Admixed American (n=1726), African ancestry (n=1550), East Asian ancestry (n=190) and South Asian ancestry (n=45). 2. AN: anorexia nervosa; BE: binge eating. Disordered eating behavior symptoms were derived from the parent-report Kiddie Schedule for Affective Disorders and Schizophrenia (KSADS). The range of AN scores 0-12; the range of BE scores 0-40. 3. Psychosocial traits (ADHD, anxiety/depression, social problems) were the summary scores of the Child Behavior Checklist (CBCL) subscales. 4. Wilcoxon rank-sum tests P values: \* p<0.05, \*\* p<0.01, \*\*\* p<0.001. The tests were performed between males and females within the study sample (i.e. EUR subgroup and Full sample respectively).

### Adult ED-PGS significantly correlated with childhood DEB

In EUR samples, AN-PGS were associated with AN symptoms in both males (AN-PGS: βmales = 0.026, 95% CI: 0.012, 0.042) and females (βfemales = 0.043, 95% CI: 0.026, 0.061). BE-PGS showed transdiagnostic effects, manifested behaviorally as AN symptoms (BE-PGS: βmales = 0.045, 95% CI: 0.030, 0.061; βfemales = 0.081, 95% CI: 0.064, 0.098) and BE symptoms (βmales = 0.034, 95% CI: 0.019, 0.050; βfemales= 0.063, 95% CI: 0.046, 0.080) (Figure **1**, see Supplementary Table 1 for estimate details). The association findings were consistent in the full sample. Sensitivity analyses using BE-NAN PGS also identified associations with AN and BE symptoms across sexes and ancestry groups (Supplementary Table 1).

**Figure 1.**
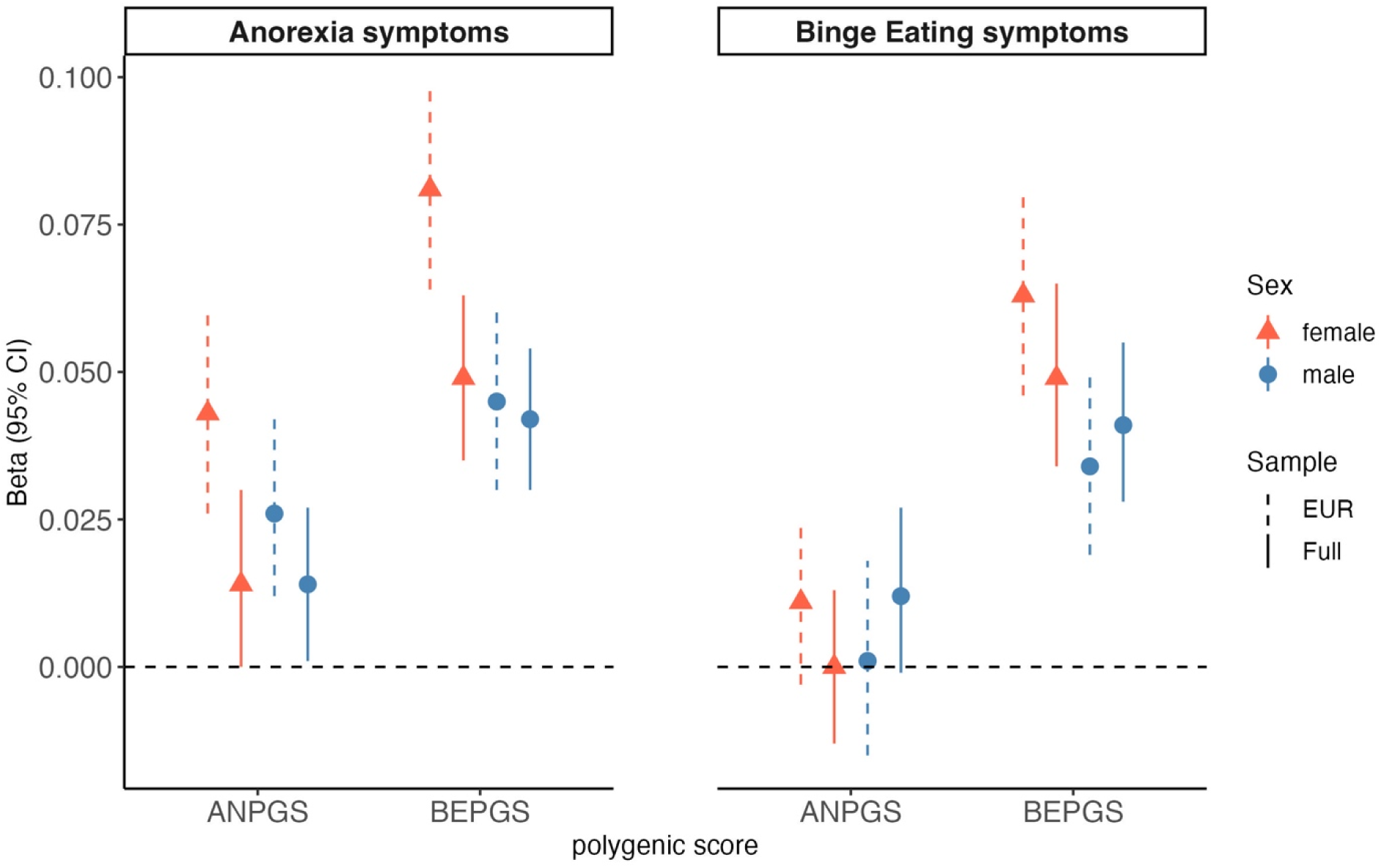
Associations between ED-PGS and childhood disordered eating behaviors. 1. AN, anorexia nervosa symptoms; ANPGS, polygenic scores for anorexia nervosa; BE, binge eating; BEPGS, polygenic scores for binge eating; EUR, European subgroup; Full, all ABCD participants (i.e. multiple genetic ancestries).

### BMI-related and psychosocial traits mediate the association between ED-PGS and childhood DEB

#### Childhood AN symptoms

Among EUR samples, negative mediation effects were found between **AN-PGS and AN symptoms** via **BMI** (ACME = -0.032, 95% CI: -0.058, -0.006), and Δ**BMI** (ACME = -0.009, 95% CI: -0.019, -0.001) **in females only**. The negative mediation resulted from sequential effects within the mediation model: higher AN-PGS were associated with lower BMI (β = −0.06, 95% CI: −0.11 to −0.01) and smaller ΔBMI (β = −0.122, 95% CI: −0.222 to −0.022), while **BMI and** Δ**BMI** positively contributed to increased AN symptoms (BMI β = 0.26, 95% CI: 0.22 to 0.29; ΔBMI β = 0.04, 95% CI: 0.02 to 0.06) (Table 2, Figure 2, Supplementary Table 2). In the full sample, BMI remained a negative mediator in females (ACME = -0.032, 95% CI: –0.051, – 0.012), while anxiety/depression emerged as a positive mediator (ACME = 0.030, 95% CI: 0.010, 0.049). No significant mediation effects were observed in males. Together, these findings suggest that the mediating mechanisms linking AN genetic risk to AN symptoms differ across sexes and possibly study sample.

**Figure 2.**
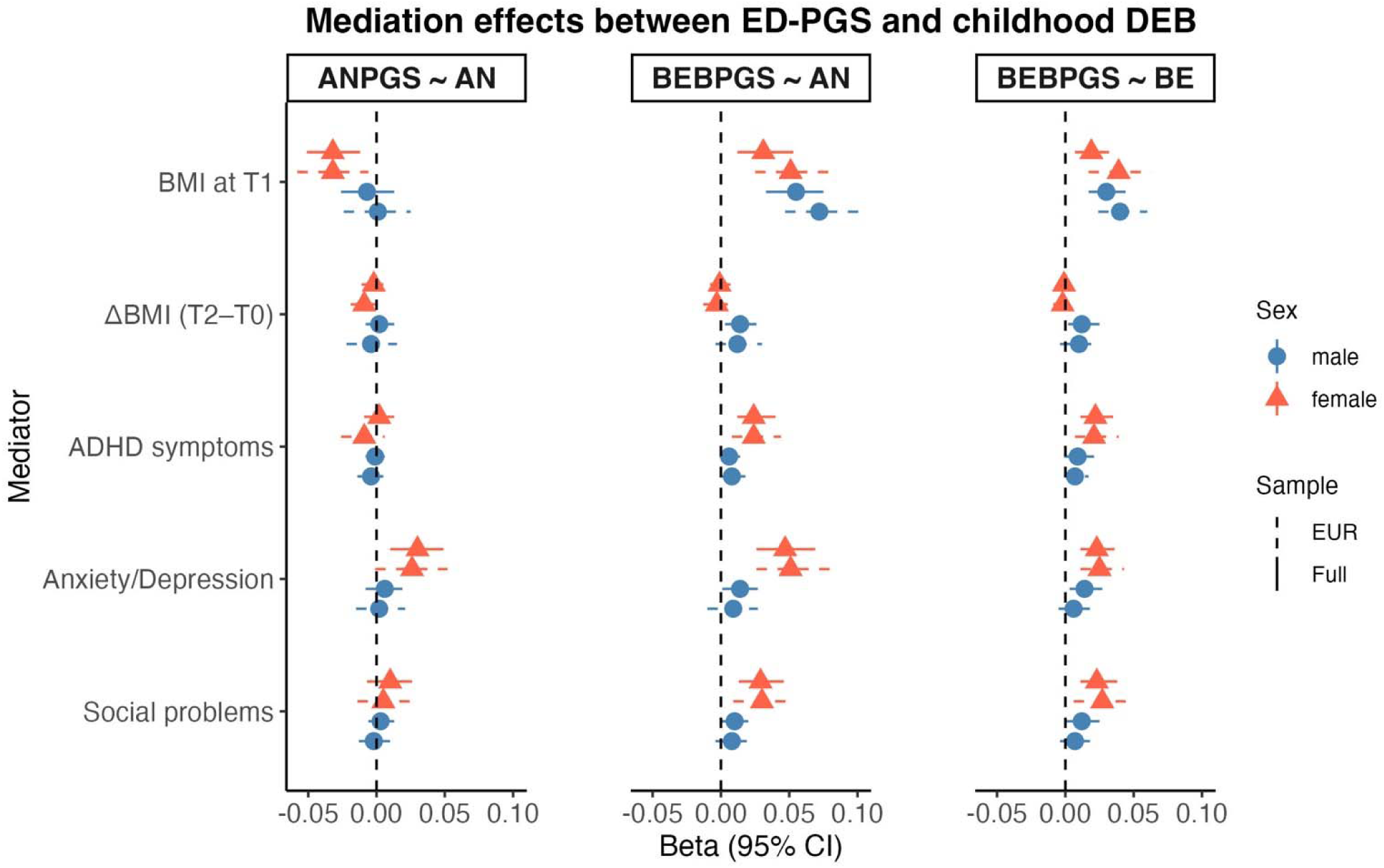
Mediation effects between ED-PGS and childhood disordered eating behaviors. 1. AN, anorexia nervosa symptoms; ANPGS, polygenic scores for anorexia nervosa; BE, binge eating; BEPGS, polygenic scores for binge eating; EUR, European subgroup; Full, all ABCD participants (i.e. multiple genetic ancestries).

**Table 2.**
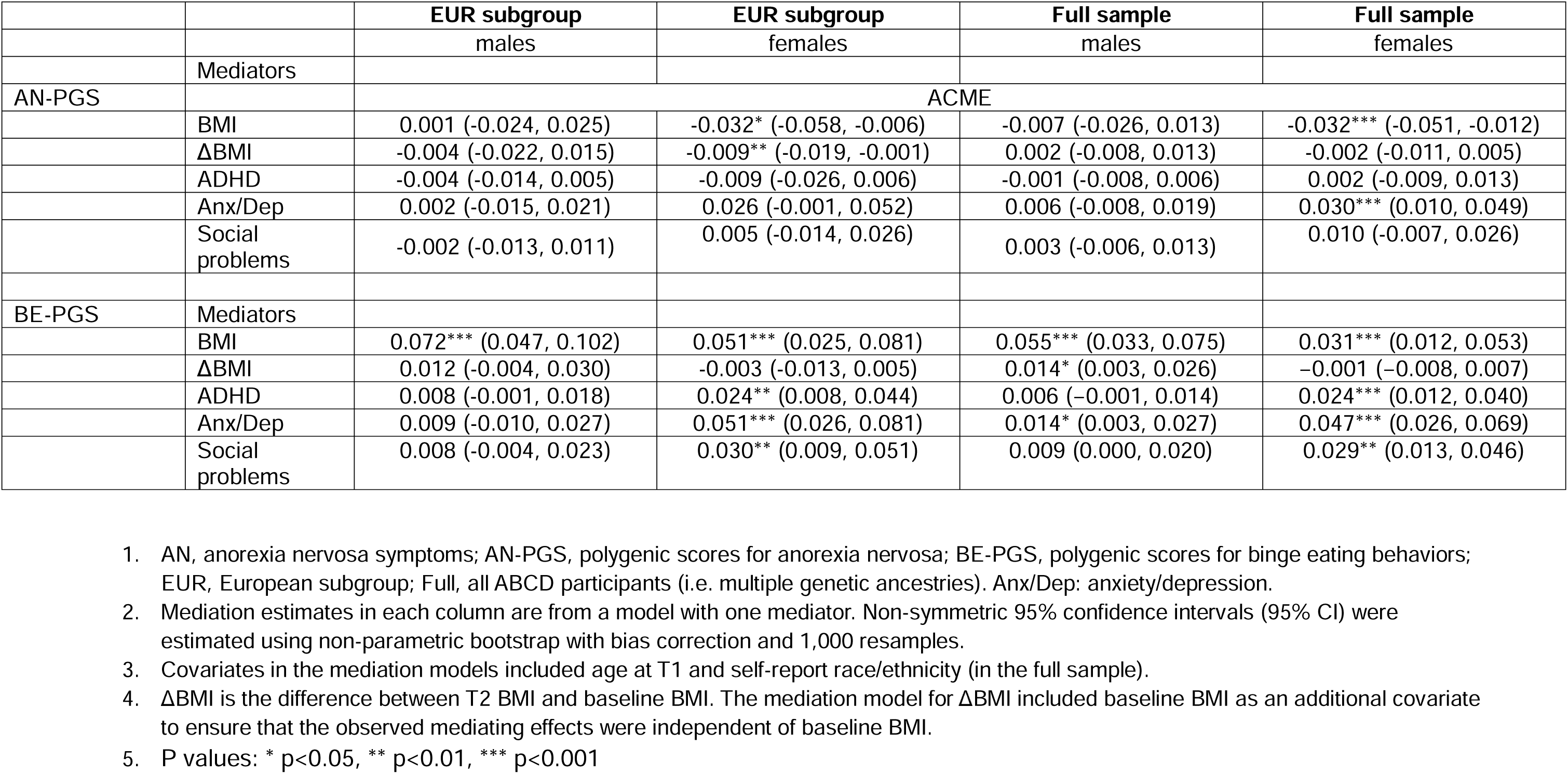
Sex-stratified Average Causal Mediation Effect estimates (ACME) between ED-PGS and AN symptoms.

For BE-PGS, multiple mediators were identified in EUR females, including BMI (ACME = 0.051, 95% CI: 0.025, 0.081), ADHD (ACME = 0.024, 95% CI: 0.008, 0.044), anxiety/depression (ACME = 0.051, 95% CI: 0.026, 0.081), and social problems (ACME = 0.030, 95% CI: 0.009, 0.051). In contrast, BMI was the sole mediator identified for EUR males (ACME = 0.072, 95% CI: 0.047, 0.102) (Table 2, Figure 2, Supplementary Table 2). These mediation patterns were largely consistent in the full sample for females. For males in the full sample, BMI (ACME = 0.055, 95% CI: 0.033, 0.075), ΔBMI (ACME = 0.014, 95% CI: 0.003, 0.026) and anxiety/depression (ACME = 0.014, 95% CI: 0.003, 0.027) were identified as mediators.

#### Childhood BE symptoms

Given the lack of significant associations between AN-PGS and BE symptoms in our mixed effect models, mediation analyses for BE symptoms focused exclusively on the relationship between BE-PGS and BE symptoms.

In EUR females, the association between BE-PGS to BE symptoms was positively mediated by BMI (ACME = 0.039, 95% CI: 0.017, 0.063), ADHD (ACME = 0.021, 95% CI: 0.007, 0.039), anxiety/depression (ACME = 0.025, 95% CI: 0.011, 0.043), and social problems (ACME = 0.027, 95% CI: 0.006, 0.049). Among EUR males, only BMI significantly mediated this association (ACME = 0.040, 95% CI: 0.024, 0.060) (Table 3, Figure 2, Supplementary Table 3). These findings were largely replicated in the full sample. Additionally, for males in the full sample, ΔBMI (ACME = 0.012, 95% CI: 0.002, 0.025) and anxiety/depression (ACME = 0.014, 95% CI: 0.003, 0.027) also emerged as significant mediators between BE-PGS and BE.

**Table 3.** Sex-stratified Average Causal Mediation Effect estimates (ACME) between ED-PGS and BE symptoms.

|  |  | **EUR subgroup** | **EUR subgroup** | **Full sample** | **Full sample** |
| --- | --- | --- | --- | --- | --- |
|  |  | males | females | males | females |
| BEB-PGS | Mediators |  |  |  |  |
|  |  | ACME |  |  |  |
|  | BMI | 0.040*** (0.024, 0.060) | 0.039*** (0.017, 0.063) | 0.030*** (0.017, 0.044) | 0.019*** (0.007, 0.032) |
| | ΔBMI | 0.010 (-0.004, 0.026) | -0.002 (-0.009, 0.004) | 0.012* (0.002, 0.025) | -0.001 (-0.006, 0.002) |
|  | ADHD | 0.007 (-0.001, 0.017) | 0.021** (0.007, 0.039) | 0.009 (-0.001, 0.021) | 0.022*** (0.011, 0.035) |
|  | Anx/Dep | 0.006 (-0.005, 0.019) | 0.025*** (0.011, 0.043) | 0.014* (0.003, 0.027) | 0.023*** (0.011, 0.036) |
|  | Social problems | 0.007 (-0.004, 0.018) | 0.027** (0.006, 0.049) | 0.012 (0.000, 0.025) | 0.023*** (0.011, 0.038) |

1. BE, binge eating; BE-PGS, polygenic scores for binge eating behaviors; EUR, European subgroup; Full, all ABCD participants (i.e. multiple genetic ancestries). Anx/Dep: anxiety/depression. 2. Mediation estimates in each column are from a model with one mediator. Non-symmetric 95% confidence intervals (95% CI) were estimated using non-parametric bootstrap with bias correction and 1,000 resamples. 3. Covariates in the mediation models included age at T1 and self-report race/ethnicity (in the full sample). 4. ΔBMI is the difference between T2 BMI and baseline BMI. The mediation model for ΔBMI included baseline BMI as an additional covariate to ensure that the observed mediating effects were independent of baseline BMI. 5. P values: \* $p < 0.05$ , \*\* $p < 0.01$ , \*\*\* $p < 0.001$

## Discussion

This study examined the associations between genetic liability for anorexia nervosa (AN) and binge eating (BE), indexed by polygenic scores (PGS), and disordered eating behaviors (DEB) across childhood and early adolescence (ages 9 to 12 years). Using summary statistics from the largest AN and BE GWAS to date, and longitudinal data from the ABCD Study, we found that higher AN-PGS was associated with increased AN symptoms while BE-PGS demonstrated transdiagnostic links to both AN and BE symptoms. These findings suggest that genetic risk factors identified in adults manifest as observable disordered eating tendencies in childhood, supporting a model of developmental continuity in ED risk. We further showed that genetic liability was partly mediated by anthropometric and psychosocial traits—including BMI, anxiety/depression, ADHD symptoms and social problems—with effects varying by sex. Identifying modifiable mediators offers key targets for early, tailored prevention strategies that address differential risk mechanisms in males and females.

Previous research has reported inconsistent associations between AN-PGS and childhood DEB. For instance, while higher AN-PGS has been linked to reduced overeating and increased undereating in childhood, it has not been shown to predict the developmental course of these behaviors ^13^. Similarly, although AN-PGS was associated with non-specific ED status in adolescent females, it was not associated with AN diagnosis ^12^. The genetic etiology of BE remains even less understood in children due to the historical lack of adequately powered GWAS, despite strong twin-based heritability estimates ^43,44^. By leveraging recent large-scale AN and BE GWAS with augmented sample sizes and enhanced statistical power, the present study provides new evidence linking AN-PGS to early AN symptoms in both sexes and demonstrates that BE-PGS shows transdiagnostic effects across AN and BE symptoms in childhood. These findings indicate that adult-derived genetic risks are manifested behaviorally in early development and can inform the identification of individuals at risk across ED phenotypes. Importantly, our findings underscore the potential of recent GWAS advances to elucidate ED risk in males, a group historically underrepresented in ED research ^2^.

A key focus of our study was to examine sex-specific mechanisms in the development of EDs. Mediation effects of BMI on the associations between AN-PGS and AN symptoms were exclusively observed in females, suggesting sex-dependent pleiotropy between the genetic risk for AN and metabolic/anthropometric traits. However, our mediation analyses revealed a paradoxical finding that the lower BMI and smaller ΔBMI predicted by higher AN-PGS served as protective factors against early AN symptoms. Specifically, lower body mass and greater weight stability were associated with a reduced burden of AN symptoms, partially offsetting the direct genetic risk indexed by AN-PGS. This pattern contrast with epidemiological evidence linking low adolescent BMI to higher AN risk in early adulthood ^45^. One explanation is that in this young community-based cohort, AN-PGS predominantly reflects the metabolic aspect of AN liability rather than its psychological or behavioral dimensions, which typically escalate from later pubertal stages into adulthood ^46^. It is possible that metabolic liability of lower BMI may become pathogenic only later, when it interacts with increasing psychological stress, body dissatisfaction, and sociocultural pressures that intensify across pubertal development. The findings regarding ΔBMI further highlight the protective effect of weight stability against eating pathology among young females during this early developmental period ^47^. Together, these findings suggest that genetically driven metabolic traits may exert shifting influences on disordered eating behaviors depending on developmental stage and co-occurring psychological factors ^30^. Further work is needed to replicate these results and clarify when and how metabolic and psychiatric components of AN risk converge to trigger the full syndrome.

Sex differences also emerged in mediation pathways for BE-PGS. Only BMI significantly mediated BE-PGS effects in male. Whereas, the effects of BE-PGS in females were additionally mediated by ADHD symptoms, anxiety/depression, and social problems—accounting for 14–29% of the associations with AN and 15–19% with BE. These findings suggest that genetic liability for BE may heighten psychosocial vulnerability in females, increasing overall susceptibility to DEBs. Importantly, these results underscore the need for a **multifaceted intervention model** that accounts for sex differences. For example, promoting healthy BMI trajectories in childhood is important for both sexes, while addressing comorbid mental health difficulties alongside monitoring physiological changes may be particularly critical for females.

This study stands among the first to demonstrate that genetic risk variants associated with AN and BE can manifest well before clinical onset of EDs. While our primary analyses focused on individuals of EUR ancestry, we observed comparable results in the diverse ABCD cohort. This suggests that cross-ancestry PGS analyses provide meaningful insights, though it should be interpreted with caution. Future GWAS that includes ancestrally diverse populations will undoubtedly improve the accuracy and generalizability of genetic risk estimates across populations. Furthermore, our longitudinal design enhances the robustness of the observed mediation pathways, providing a clearer temporal framework for how genetic risk manifests in early development.

Despite these strengths, several limitations warrant consideration. First, we used KSADS-ED module items to assess DEB. While these items align with clinical diagnostic criteria, they may not capture subthreshold or prodromal ED behaviors such as loss of control eating, potentially biasing the sample toward more severe cases. As such, our findings may not generalize to milder forms of DEB and may underestimate the association between genetic liability and broader disordered eating symptoms. Future research should incorporate developmentally sensitive and dimensional phenotypes. Second, we were unable to assess subjective experiences such as weight stigma and body dissatisfaction, which are known predictors of EDs ^48,49^ and could not be captured by objective weight status alone. Including these constructs in future studies would yield a more comprehensive understanding of ED risk pathways. Third, while some sample overlap between AN and BE GWAS may have contributed to shared PGS effects, sensitivity analyses using BE GWAS excluding AN-ascertained cohorts yielded consistent results. Moreover, despite the presence of BE symptoms within the AN GWAS samples, AN-PGS showed no significant association with childhood BE symptoms. Together, these results suggest that the observed associations likely reflect the heterogeneous nature of childhood DEB.

In conclusion, our study demonstrates that genetic liabilities to EDs manifest early and through sex-dependent pathways. While AN-PGS showed specificity for childhood AN symptoms, BE-PGS may serve as a broader indicator of early ED liability. The observed mediation effects suggest that genetic influences operate partly through modifiable developmental processes and highlight actionable intervention points—physiological (e.g., healthy BMI trajectories), psychological (e.g., mood regulation, impulsivity), and social (e.g., peer relationships) domains. Altogether, these findings underscore the potential of integrating genetic liability markers (e.g. PGS) with environmental and psychosocial factors to identify high-risk individuals and specific vulnerabilities to develop tailored, multifaceted interventions during critical developmental windows.

## Supporting information

Eating Disorders Working Group of the Psychiatric Genomics Consortium

## Data Availability

All data produced in the present study are available upon reasonable request to the authors.

## Acknowledgement

Data used in the preparation of this article were obtained from the Adolescent Brain Cognitive Development^SM^ (ABCD) Study (https://abcdstudy.org), held in the NIMH Data Archive (NDA). This is a multisite, longitudinal study designed to recruit more than 10,000 children age 9-10 and follow them over 10 years into early adulthood. The ABCD Study® is supported by the National Institutes of Health and additional federal partners under award numbers U01DA041048, U01DA050989, U01DA051016, U01DA041022, U01DA051018, U01DA051037, U01DA050987, U01DA041174, U01DA041106, U01DA041117, U01DA041028, U01DA041134, U01DA050988, U01DA051039, U01DA041156, U01DA041025, U01DA041120, U01DA051038, U01DA041148, U01DA041093, U01DA041089, U24DA041123, U24DA041147. A full list of supporters is available at https://abcdstudy.org/federal-partners.html. A listing of participating sites and a complete listing of the study investigators can be found at https://abcdstudy.org/consortium_members/. ABCD consortium investigators designed and implemented the study and/or provided data but did not necessarily participate in the analysis or writing of this report. This manuscript reflects the views of the authors and may not reflect the opinions or views of the NIH or ABCD consortium investigators.

The ABCD data repository grows and changes over time. The ABCD data used in this report came from NIMH Data Archive Digital Object Identifier (DOI) <u>10.15154/z563-zd24</u>. DOIs can be found at http://dx.doi.org/10.15154/z563-zd24.

