## supplementary tables and figures for "Childhood Mental Health and Body Mass Index as Mediators of Genetic Risk for Eating Disorders"

Supplementary Table 1. Associations between ED-PGS and childhood disordered eating behaviors

|  |  | **EUR subgroup** | | | | **Full sample** | | | |
| --- | --- | --- | --- | --- | --- | --- | --- | --- | --- |
|  |  | **Male** |  | **Female** |  | **Male** |  | **Female** |  |
| **PGS** | **Trait** | **beta** | **R^2^** | **beta** | **R^2^** | **beta** | **R^2^** | **beta** | **R^2^** |
| AN | AN | 0.026 (0.012, 0.042) ** | 0.003 | 0.043 (0.026, 0.061) *** | 0.003 | 0.014 (0.001, 0.027)* | 0.010 | 0.014 (0.000, 0.030) * | 0.006 |
| BE |  | 0.045 (0.030, 0.061) *** | 0.004 | 0.081 (0.064, 0.098) *** | 0.007 | 0.042 (0.030, 0.054) *** | 0.011 | 0.049 (0.035, 0.063) *** | 0.008 |
| BE-NAN |  | 0.039 (0.022, 0.054) *** | 0.004 | 0.073 (0.057, 0.089) *** | 0.006 | 0.052 (0.040, 0.064) *** | 0.012 | 0.055 (0.042, 0.069) *** | 0.008 |
| BMI |  | 0.097 (0.082, 0.112) *** | 0.013 | 0.150 (0.133, 0.167) *** | 0.024 | 0.103 (0.090, 0.115) *** | 0.020 | 0.112 (0.097, 0.127) *** | 0.017 |
| AN | BE | 0.001 (-0.015, 0.018) | 0.000 | 0.011 (-0.003, 0.025) | 0.001 | 0.012 (-0.001, 0.027) | 0.004 | 0.000 (-0.013, 0.013) | 0.002 |
| BE |  | 0.034 (0.019, 0.050) *** | 0.001 | 0.063 (0.046, 0.080) *** | 0.004 | 0.041 (0.028, 0.055) *** | 0.006 | 0.049 (0.034, 0.065) *** | 0.004 |
| BE-NAN |  | 0.035 (0.018, 0.050) *** | 0.002 | 0.066 (0.049, 0.082) *** | 0.005 | 0.045 (0.030, 0.060) *** | 0.006 | 0.060 (0.043, 0.076) *** | 0.005 |
| BMI |  | 0.100 (0.077, 0.118) *** | 0.012 | 0.113 (0.091, 0.133) *** | 0.014 | 0.099 (0.080, 0.114) *** | 0.014 | 0.090 (0.073, 0.106) *** | 0.009 |

1. Estimates of the mixed effect model with random intercept. ED-PGS were adjusted for genetic PCs and genotyping plates before entering the regression models. The final models control for covariates including concurrent age in the EUR models, and age and self-reported race ethnicity in the full sample.
2. FDR corrected P values: * p<0.05, ** p<0.01, *** p<0.001
3. R^2^: fixed effect R-square values in the mixed effect model.
4. BE-NAN: BE not-ascertained-for-AN. BE-NAN GWAS was performed in a subset of samples of BE-BROAD GWAS, which excluded cases recruited through studies that focused on AN ascertainment to understand the influence of AN on BE-BROAD phenotype.

Supplementary Table 2. Estimates of sex-stratified mediation model between ED-PGS and AN symptoms

| AN-PGS | | | | | | |
| --- | --- | --- | --- | --- | --- | --- |
| Sample | estimates | Mediator | | | | |
|  |  | BMI (95% CI) | ΔBMI (95% CI) | ADHD (95% CI) | Anx/Dep (95% CI) | Social (95% CI) |
| EUR Male | ACME | 0.001 (-0.024, 0.025) | -0.004 (-0.022, 0.015) | -0.004 (-0.014, 0.005) | 0.002 (-0.015, 0.021) | -0.002 (-0.013, 0.011) |
|  | ADE | 0.097 (0.024, 0.170) | 0.101 (0.030, 0.170) | 0.100 (0.020, 0.176) | 0.094 (0.020, 0.169) | 0.098 (0.017, 0.175) |
|  | Total | 0.098 (0.028, 0.175) | 0.097 (0.022, 0.168) | 0.096 (0.018, 0.171) | 0.096 (0.022, 0.176) | 0.096 (0.016, 0.178) |
|  | % mediated |  |  |  |  |  |
| EUR Female | ACME | -0.032* (-0.058, -0.006) | -0.009** (-0.019, -0.001) | -0.009 (-0.026, 0.006) | 0.026 (-0.001, 0.052) | 0.005 (-0.014, 0.026) |
|  | ADE | 0.094 (0.013, 0.182) | 0.078 (-0.007, 0.168) | 0.085 (-0.001, 0.175) | 0.049 (-0.034, 0.135) | 0.070 (-0.016, 0.154) |
|  | Total | 0.062 (-0.026, 0.152) | 0.069 (-0.016, 0.162) | 0.075 (-0.008, 0.164) | 0.075 (-0.009, 0.165) | 0.075 (-0.014, 0.160) |
|  | % mediated | N/A^$^ | N/A^$^ |  |  |  |
| Full sample Male | ACME | -0.007 (-0.026, 0.013) | 0.002 (-0.008, 0.013) | -0.001 (-0.008, 0.006) | 0.006 (-0.008, 0.019) | 0.003 (-0.006, 0.013) |
|  | ADE | 0.043 (-0.019, 0.100) | 0.031 (-0.026, 0.094) | 0.033 (-0.030, 0.096) | 0.026 (-0.029, 0.083) | 0.029 (-0.038, 0.091) |
|  | Total | 0.037 (-0.030, 0.096) | 0.033 (-0.024, 0.098) | 0.032 (-0.031, 0.093) | 0.032 (-0.026, 0.091) | 0.032 (-0.036, 0.096) |
|  | % mediated |  |  |  |  |  |
| Full sample Female | ACME | -0.032*** (-0.051, -0.012) | -0.002 (-0.011, 0.005) | 0.002 (-0.009, 0.013) | 0.030*** (0.010, 0.049) | 0.010 (-0.007, 0.026) |
|  | ADE | 0.071 (-0.001, 0.139) | 0.069 (-0.004, 0.145) | 0.051 (-0.023, 0.122) | 0.022 (-0.048, 0.087) | 0.043 (-0.027, 0.115) |
|  | Total | 0.039 (-0.035, 0.107) | 0.067 (-0.005, 0.141) | 0.052 (-0.020, 0.122) | 0.052 (-0.018, 0.125) | 0.052 (-0.017, 0.125) |
|  | % mediated | N/A^$^ |  |  | 58% |  |
| BEB-PGS | | | | | | |
| Sample | estimates | Mediator | | | | |
|  |  | BMI (95% CI) | ΔBMI (95% CI) | ADHD (95% CI) | Anx/Dep (95% CI) | Social (95% CI) |
| EUR Male | ACME | 0.072*** (0.047, 0.102) | 0.012 (-0.004, 0.030) | 0.008 (-0.001, 0.018) | 0.009 (-0.010, 0.027) | 0.008 (-0.004, 0.023) |
|  | ADE | 0.087 (0.008, 0.173) | 0.046 (-0.031, 0.128) | 0.151 (0.071, 0.234) | 0.150 (0.070, 0.234) | 0.151 (0.071, 0.234) |
|  | Total | 0.159 (0.078, 0.253) | 0.058 (-0.020, 0.142) | 0.159 (0.077, 0.244) | 0.159 (0.074, 0.243) | 0.159 (0.078, 0.242) |
|  | % mediated | 45 |  |  |  |  |
| EUR Female | ACME | 0.051*** (0.025, 0.081) | -0.003 (-0.013, 0.005) | 0.024** (0.008, 0.044) | 0.051*** (0.026, 0.081) | 0.030** (0.009, 0.051) |
|  | ADE | 0.122 (0.040, 0.205) | 0.002 (0.179, 0.128) | 0.153 (0.072, 0.235) | 0.126 (0.052, 0.207) | 0.148 (0.064, 0.231) |
|  | Total | 0.173 (0.086, 0.261) | -0.001 (0.175, 0.142) | 0.177 (0.101, 0.260) | 0.177 (0.089, 0.258) | 0.177 (0.095, 0.258) |
|  | % mediated | 29 |  | 14 | 29 | 17 |
| Full sample Male | ACME | 0.055*** (0.033, 0.075) | 0.014* (0.003, 0.026) | 0.006 (−0.001, 0.014) | 0.014* (0.001, 0.027) | 0.009 (0.000, 0.020) |
|  | ADE | 0.041 (−0.026, 0.109) | 0.015 (−0.051, 0.082) | 0.089 (0.024, 0.158) | 0.081 (0.014, 0.146) | 0.086 (0.019, 0.151) |
|  | Total | 0.096 (0.026, 0.167) | 0.028 (−0.039, 0.094) | 0.096 (0.033, 0.162) | 0.096 (0.029, 0.157) | 0.096 (0.029, 0.162) |
|  | % mediated | 57 | 50 |  | 15 |  |
| Full sample Female | ACME | 0.031*** (0.012, 0.053) | −0.001 (−0.008, 0.007) | 0.024*** (0.012, 0.040) | 0.047*** (0.026, 0.069) | 0.029** (0.013, 0.046) |
|  | ADE | 0.101 (0.037, 0.171) | 0.072 (−0.002, 0.145) | 0.118 (0.049, 0.188) | 0.098 (0.034, 0.171) | 0.113 (0.038, 0.183) |
|  | Total | 0.132 (0.064, 0.203) | 0.071 (−0.002, 0.145) | 0.142 (0.067, 0.212) | 0.142 (0.071, 0.215) | 0.142 (0.072, 0.210) |
|  | % mediated | 23 |  | 17 | 33 | 20 |

1. ACME, average causal mediation effect; ADE, average direct effect; % mediated= mediated effect/total effect
2. ^$^ : NA, proportion mediated is not applicable when total effect and mediated effect are in the opposite direction. BMI and ΔBMI negatively mediated the association between AN-PGS and AN symptoms in females.
3. Confidence intervals were derived from non-parametric bootstrapping with 1,000 simulations.
4. P values: * p<0.05, ** p<0.01, *** p<0.001.

Supplementary Table 3. Estimates of sex-stratified mediation model between ED-PGS and BE symptoms

| BEB-PGS | | | | | | |
| --- | --- | --- | --- | --- | --- | --- |
| Sex | estimates | Mediator | | | | |
|  |  | BMI (95% CI) | ΔBMI (95% CI) | ADHD (95% CI) | Anx/Dep (95% CI) | Social (95% CI) |
| EUR Male | ACME | 0.040*** (0.024, 0.060) | 0.010 (-0.004, 0.026) | 0.007 (-0.001, 0.017) | 0.006 (-0.005, 0.019) | 0.007 (-0.004, 0.018) |
|  | ADE | 0.053 (-0.008, 0.113) | 0.032 (-0.030, 0.084) | 0.083 (0.022, 0.145) | 0.085 (0.022, 0.145) | 0.083 (0.020, 0.148) |
|  | Total | 0.094 (0.027, 0.160) | 0.042 (-0.021, 0.097) | 0.090 (0.027, 0.156) | 0.090 (0.025, 0.149) | 0.090 (0.026, 0.156) |
|  | % mediated | 43 |  |  |  |  |
| EUR Female | ACME | 0.039*** (0.017, 0.063) | -0.002 (-0.009, 0.004) | 0.021** (0.007, 0.039) | 0.025*** (0.011, 0.043) | 0.027** (0.006, 0.049) |
|  | ADE | 0.094 (0.020, 0.169) | 0.057 (-0.008, 0.120) | 0.120 (0.047, 0.203) | 0.116 (0.042, 0.196) | 0.114 (0.040, 0.190) |
|  | Total | 0.132 (0.058, 0.213) | 0.055 (-0.010, 0.121) | 0.141 (0.069, 0.228) | 0.141 (0.064, 0.223) | 0.141 (0.070, 0.223) |
|  | % mediated | 30 |  | 15 | 18 | 19 |
| Full sample Male | ACME | 0.030*** (0.017, 0.044) | 0.012* (0.002, 0.025) | 0.009 (-0.001, 0.021) | 0.014* (0.003, 0.027) | 0.012 (0.000, 0.025) |
|  | ADE | 0.060 (-0.000, 0.118) | 0.026 (-0.039, 0.083) | 0.084 (0.017, 0.146) | 0.080 (0.016, 0.142) | 0.086 (0.021, 0.155) |
|  | Total | 0.090 (0.029, 0.148) | 0.038 (-0.026, 0.096) | 0.094 (0.027, 0.159) | 0.094 (0.029, 0.157) | 0.094 (0.033, 0.159) |
|  | % mediated | 33 | 32 |  | 15 |  |
| Full sample Female | ACME | 0.019*** (0.007, 0.032) | -0.001 (-0.006, 0.002) | 0.022*** (0.011, 0.035) | 0.023*** (0.011, 0.036) | 0.023*** (0.011, 0.038) |
|  | ADE | 0.083 (0.011, 0.152) | 0.062 (-0.015, 0.137) | 0.084 (0.012, 0.157) | 0.084 (0.015, 0.159) | 0.084 (0.008, 0.152) |
|  | Total | 0.102 (0.029, 0.174) | 0.061 (-0.018, 0.137) | 0.106 (0.037, 0.186) | 0.106 (0.037, 0.186) | 0.106 (0.029, 0.174) |
|  | % mediated | 19 |  | 21 | 22 | 22 |

1. ACME, average causal mediation effect; ADE, average direct effect; % mediated= mediated effect/total effect.
2. Confidence intervals were derived from non-parametric bootstrapping with 1,000 simulations.
3. P values: * p<0.05, ** p<0.01, *** p<0.001.

Supplementary Figure 1. Distribution of DEB scores

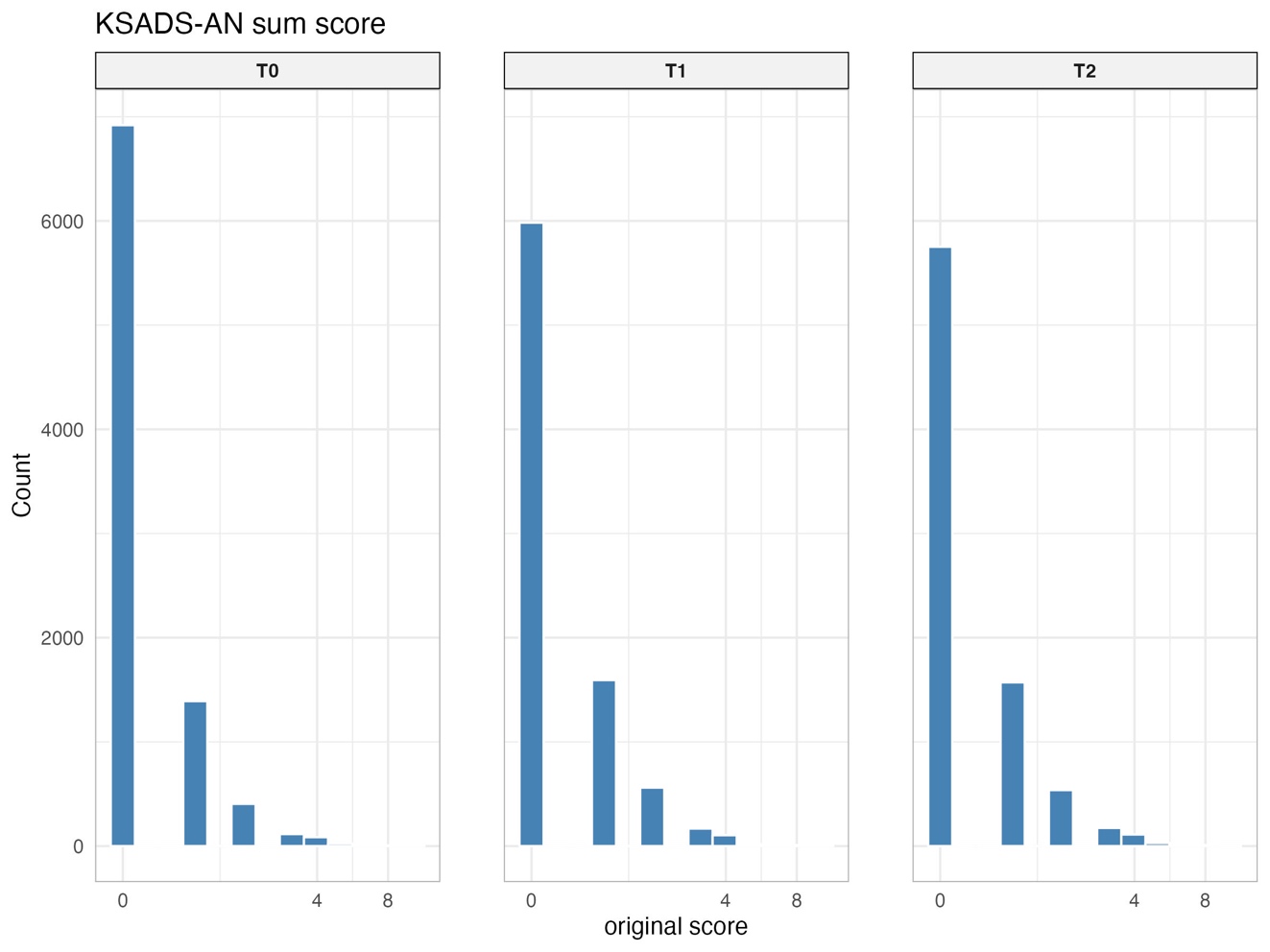

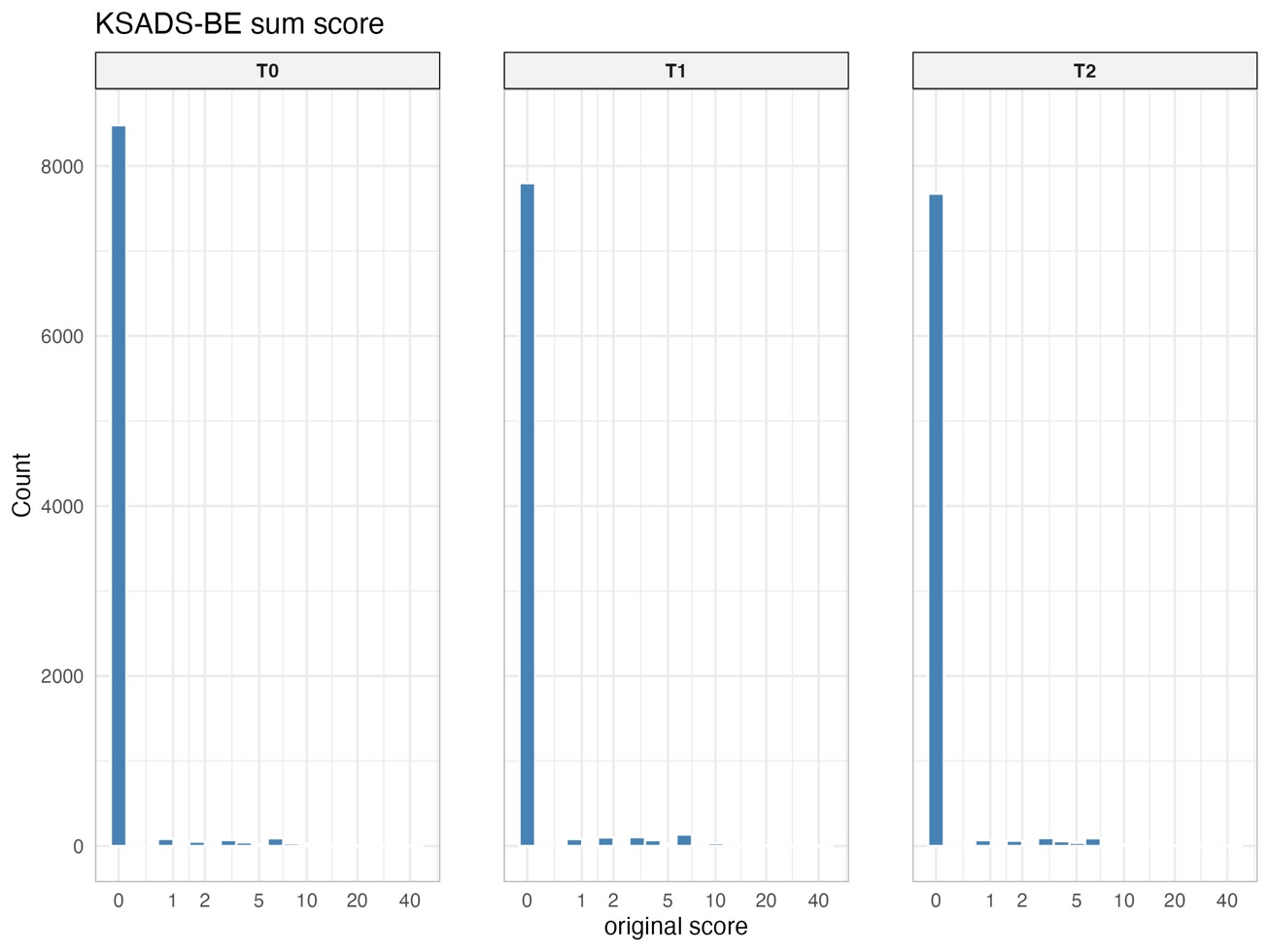

AN and BE symptom scores were derived from the parent-report Kiddie Schedule for Affective Disorders and Schizophrenia (KSADS). The range of AN scores is 0-12. The range of BE scores 0-40.
