## Supplementary material for "Childhood Mental Health and Body Mass Index as Mediators of Genetic Risk for Eating Disorders": Eating Disorders Working Group of the Psychiatric Genomics Consortium

Dr. Jet D Termorshuizen PhD^1^; Dr. Helena L Davies PhD^2,3,4^; Mr. Sang-Hyuck Lee MSc^2,5^; Dr. Jessica K Dennis PhD^6,7^; Dr. Christopher Hübel MD, PhD^8,9,2^; Ms. Jessica S Johnson MPH, MFA^10^; Dr. Yi Lu PhD^1^; Dr. Melissa A Munn-Chernoff PhD^11^; Dr. Triinu Peters PhD^12,13,14^; Baiyu Qi MPH^15^; Dr. Katherine E Schaumberg PhD^16,17^; Dr. Rebecca H Signer MS, PhD^18^; Karanvir Singh MSc^7^; Dr. Abigail R ter Kuile PhD^19,2,5^; Dr. Laura M Thornton PhD^10^; Dr. Jiayi Xu PhD^20^; Dr. Shuyang Yao PhD^1^; Dr. Zeynep Yilmaz PhD^8,21,1,10^; Dr. Ruyue Zhang PhD^1,22^; Dr. Johan Zvrskovec PhD^2,23^; Dr. Mohamed Abdulkadir PhD^8^; Dr. Ziada Ayorech PhD^24^; Dr. Elizabeth C Corfield^25,26,27^; Dr. Alexandra Havdahl PhD^25,26,24^; Dr. Kristi Krebs PhD^28^; Taralynn M Mack^29^; Dr. Maria Niarchou PhD^30^; Teemu Palviainen^31^; Dr. Julia M Sealock PhD^32,33^; Dr. Jessica H Baker PhD^34^; Dr. Andrew W Bergen PhD^35,36^; Dr. Andreas Birgegård PhD^1^; Prof. Vesna Boraska Perica PhD^37^; Dr. Katharina Bühren MD^38^; Dr. Roland Burghardt MD^39^; Prof. Matteo Cassina MD^40^; Dr. Giovanni Castellini PhD, MD^155^; Dr. Enrico Collantoni MD, PhD^41^; Dr. James J Crowley PhD^22,42^; Dr. Unna N Danner PhD^43^; Dr. Franziska Degenhardt MD^44^; Prof. Janiece E DeSocio PhD, RN, PMHNP-BC^45^; Dr. Christian Dina PhD^46^; Prof Monika Dmitrzak-Węglarz PhD^47^; Dr. Laramie E Duncan PhD^48^; Dr. Karin M Egberts MD, PhD^49,50^; Dr. Lenka Foretova MD, PhD^51^; Dr. Ina Giegling PhD^52^; Dr. Fragiskos Gonidakis MD^53^; Dr. Scott D Gordon PhD^54^; Dr. Jakob Grove PhD^21,55,56,57^; Prof. Sébastien Guillaume MD, PhD^58^; Dr. Jerry D Guintivano PhD^10,22^; Dr. Annette M Hartmann MD^52^; Dr. Konstantinos Hatzikotoulas PhD^59^; Mr. Stefan Herms MSc^60,61,62^; Dr. Hartmut Imgart MD^63^; Dr. Susana Jiménez-Murcia PhD^64,65,66,67,68^; Dr. Antonio Julià PhD^69^; Dr. Gursharan Kalsi PhD^2^; Dr. Deborah Kaminská PhD^70^; Adjunct Professor Leila J Karhunen PhD^71^; Dr. Hannah L. Kennedy PhD^129^; Dr. Kirsty M Kiezebrink PhD, FHEA, RNutr^72^; Theresa Kolb MSc^73,74^; Dr. Janne T Larsen PhD^8,55^; Dr. Dong Li PhD^75,76,77^; Dr. Lisa Lilenfeld PhD^78^; Prof. Mario Maj MD, PhD^79^; Dr. Morten Mattingsdal PhD^80,81^; Dr. Paolo Meneguzzo MD; PhD^41,82^; Ms. Allison L Miller PGDipSc^83^; Dr. Karen S Mitchell PhD^84,85^; Prof. Alessio Maria Monteleone MD, PhD^86^; Dr. Catherine M Olsen PhD, MPH^87^; Dr. Leonid Padyukov MD, PhD^88^; Mr. Richard Parker BA(Hons)^54^; Dr. Michaela A. Pettie PhD^83^; Dr. Dalila Pinto PhD^90,91^; Dr. Anu Raevuori MD, PhD^92,93^; Prof. Samuli Ripatti PhD^31,93,32^; Dr. Marion E Roberts PhD^94,95^; Prof. Paolo Santonastaso MD^41^; Androula Savva MSc^42^; Prof. Ulrike H Schmidt MD, PhD^95^; Prof. Alexandra Schosser MD, PhD, MBA^96^; Prof. Jochen Seitz MD^44,14^; Dr. Lenka LS Slachtova PhD^97^; Prof. Agnieszka Slopien MD, PhD^98^; Prof. Sandro Sorbi MD^99^; Peter S Straub MS^30^; Dr. Jin P Szatkiewicz PhD^22^; Dr. Friederike I Tam MD^73^; Prof. Elena Tenconi PsyD, PhD^41,82^; Prof. Alfonso Tortorella MD^100^; Prof. Artemis Tsitsika MD, PhD^101^; Prof. Annemarie A van Elburg MD, PhD^43,102^; Dr. Gudrun Wagner Dr, MSc, DPO^103^; Dr. Hunna J Watson PhD, MPsychClin, MBiostats^10,104^; Prof. Roger AH Adan PhD^105,43,106^; Prof. Lars Alfredsson PhD^107,108^; Prof. Ole A Andreassen MD, PhD^109,81,110^; Prof. Helga Ask PhD^25,24^; Dr. Anders D. Børglum MD, PhD^21,55,56^; Dr. Harry A Brandt MD^111,112^; Dr. Steven Crawford MD^112^; Prof. Scott Crow MD^113^; Dr. Lea K Davis PhD^18,114,115^; Prof. Martina de Zwaan MD^116^; Prof. George Dedoussis PhD^117^; Dr. Danielle M Dick PhD^118^; Prof. Stefan Ehrlich MD, PhD^73^; Prof. Xavier Estivill MD, PhD^120^; Prof. Angela Favaro MD, PhD^41,82^; Prof. Fernando Fernández-Aranda PhD^64,65,67,66^; Dr. Krista Fischer PhD^121,28^; Prof. Dr. Andreas J Forstner MD^62,122,123^; Prof. Philip Gorwood MD, PhD^124,125^; Prof. Hakon Hakonarson MD, PhD^75,77^; Prof. Johannes Hebebrand MD^44^; Prof. Beate Herpertz-Dahlmann MD^126^; Prof. Anke Hinney PhD^12,13^; Prof. James I Hudson MD, ScD^127^; Dr. Craig Johnson PhD^128^; Dr. Jennifer Jordan PhD^129,130^; Prof. Allan S Kaplan MD^131^; Prof. Jaakko Kaprio MD, PhD^31^; Prof. Andreas FK Karwautz MD^132^; Prof. Martien JH Kas PhD^105,133^; Prof. Walter H Kaye MD^134^; Prof. James L Kennedy MD, FRCP(C)^135,136^; Prof. Martin A Kennedy PhD^83^; Dr. Anna Keski-Rahkonen MD, PhD, MPH^93^; Prof. Youl-Ri Kim MD, PhD^137^; Prof. Kelly L Klump PhD^138^; Dr. Mikael Landén MD, PhD^139,1^; Prof. Stéphanie Le Hellard PhD^140^; Dr. Kelli Lehto PhD^28^; Dr. Qingqin S. Li PhD^171^; Prof. Jolanta Lissowska PhD^141^; Dr. Jurjen J. Luykx MD,PhD^172,173,174^; Prof. Sarah L Maguire BScPsych(Hons), M.A., DCP, PhD^142^; Prof. Nicholas G Martin PhD^54^; Prof. Manuel Mattheisen MD^143,144^; Prof. Sarah E Medland PhD^145,146,147^; Prof. Philip Mehler MD, FACP, FAED, CEDS^175,128^; Prof. Nadia Micali MD, PhD^3,148,4^; Prof. James E Mitchell MD^149^; Prof. Palmiero Monteleone MD^150^; Prof. Preben Bo Mortensen MD, DrMedSc^8^; Prof. Benedetta Nacmias PhD^99^; Prof. Roel A Ophoff PhD^151^; Prof. Hana Papezova MD, PhD, FAED^70^; Prof. Nancy L Pedersen PhD^1^; Dr. Liselotte V Petersen PhD^8,55^; Dr. Louisa S Rajcsanyi PhD^12,13^; Dr. Nicolas Ramoz PhD^152,153^; Prof. Ted Reichborn-Kjennerud MD, PhD^154,25^; Prof. Valdo Ricca MD^155^; Dr. Stephan Ripke MD, PhD^156,33,157^; Prof. Dan Rujescu MD^52^; Prof. Filip Rybakowski MD, PhD^158^; Prof. Stephen W Scherer PhD, FRSC^159,160^; Dr. Margarita CT Slof-Op 't Landt PhD^161,162^; Prof. Howard Steiger, PhD^169,170^, Prof. Patrick F Sullivan MD, FRANZCP^22,10^; Dr. Beata Świątkowska PhD^163^; Prof. Eric F van Furth PhD^161^; Prof. Tracey D Wade PhD^164^; Prof. Thomas Werge PhD^165,4^; Prof. David C Whiteman MBBS(Hons), PhD, FAFPHM^87^; Prof. D. Blake Woodside MD, MSc, FRCPC^135^; Dr. Ya-Ke Wu, PhD, RN^89,10^; Prof. Stephan Zipfel MD^166,167^; Eating Disorders Working Group of the Psychiatric Genomics Consortium; Estonian Biobank (EstBB); Prof. Cynthia M Bulik PhD^10,1,168^; Dr. Laura M Huckins PhD^20^; Prof. Gerome Breen PhD^2,5^; Dr. Jonathan RI Coleman PhD^2,5^

1	Department of Medical Epidemiology and Biostatistics; Karolinska Institutet; Stockholm; Sweden

2	Institute of Psychiatry, Psychology and Neuroscience, Social, Genetic and Developmental Psychiatry (SGDP) Centre; King's College London; London; United Kingdom

3	Center for Eating and feeding Disorders Research, Mental Health Center Ballerup; Copenhagen University Hospital - Mental Health Services; Copenhagen; Denmark

4	Institute of Biological Psychiatry; Mental Health Center Sct. Hans; Mental Health Services Copenhagen; Roskilde; Denmark

5	National Institute for Health Research Biomedical Research Centre; King's College London and South London and Maudsley National Health Service Trust; London; United Kingdom

6	Department of Medical Genetics; University of British Columbia; Vancouver; British Columbia; Canada

7	Graduate Program in Bioinformatics; University of British Columbia; Vancouver; British Columbia; Canada

8	National Centre for Register-based Research; Aarhus University; Aarhus; Denmark

9	Clinic for Child and Adolescent Psychiatry, Psychotherapy and Psychosomatics; German Red Cross Hospitals Berlin; Berlin; Germany

10	Department of Psychiatry; University of North Carolina at Chapel Hill; Chapel Hill; North Carolina; United States

11	Department of Community, Family, and Addiction Sciences; Texas Tech University; Lubbock; Texas; United States

12	Section for Molecular Genetics in Mental Disorders; LVR University Clinic Essen, University of Duisburg-Essen; Essen; Northrhine-Westfalia; Germany

13	Institute of Sex and Gender-Sensitive Medicine; University Hospital Essen, University of Duisburg-Essen; Essen; Northrhine-Westfalia; Germany

14	Center for Translational Neuro- and Behavioral Sciences; University Hospital Essen, University of Duisburg-Essen; Essen; Northrhine-Westfalia; Germany

15	Department of Epidemiology; University of North Carolina at Chapel Hill; Chapel Hill; North Carolina; United States

16	Department of Psychiatry; University of Wisconsin; Madison; Wisconsin; United States

17	Department of Psychology; University of Texas; Austin; Texas; United States

18	Department of Genetics and Genomic Sciences; Icahn School of Medicine at Mount Sinai; New York; New York; United States

19	Department of Clinical, Educational, and Health Psychology; University College London; London; United Kingdom

20	Department of Psychiatry; Yale University; New Haven; Connecticut; United States

21	Department of Biomedicine; Aarhus University; Aarhus; Denmark

22	Department of Genetics; University of North Carolina at Chapel Hill; Chapel Hill; North Carolina; United States

23	National Institute for Health and Care Research (NIHR) Maudsley Biomedical Research Centre; South London and Maudsley NHS Foundation Trust; London; United Kingdom

24	Department of Psychology; PROMENTA Research Centre; University of Oslo; Oslo; Norway

25	PsychGen Centre for Genetic Epidemiology and Mental Health; Norwegian Institute of Public Health; Oslo; Norway

26	Psychiatric Genetic Epidemiology Group, Research Department; Lovisenberg Diakonale Hospital; Oslo; Norway

27	MRC Integrative Epidemiology Unit, Population Health Sciences; Bristol Medical School; University of Bristol; Bristol; United Kingdom

28	Estonian Genome Centre, Institute of Genomics; University of Tartu; Tartu; Estonia

29	Vanderbilt Genetics Institute; Vanderbilt University; Nashville; Tennessee; United States

30	Department of Genetic Medicine; Vanderbilt University Medical Center; Nashville; Tennessee; United States

31	Institute for Molecular Medicine Finland FIMM, Helsinki Institute of Life Science HiLIFE; University of Helsinki; Helsinki; Finland

32	Analytic and Translational Genetics Unit; Broad Institute of the Massachusetts Institute of Technology and Harvard University; Massachusetts General Hospital; Boston; Massachusetts; United States

33	Stanley Center for Psychiatric Research; Broad Institute of the Massachusetts Institute of Technology and Harvard University; Cambridge; Massachusetts; United States

34	Independent Researcher; Mebane; North Carolina; United States

35	Oregon Research Institute; Springfield; Oregon; United States

36	Department of Medicine; New Jersey Medical School, Rutgers University; Newark; New Jersey; United States

37	Department for Medical Biology; University of Split School of Medicine; Split; Croatia

38	Department of Child and Adolescent Psychiatry, Psychosomatics and Psychotherapy; Ludwig-Maximilians-Universität München; Munich; Germany

39	Department of Child and Adolescent Psychiatry; Oberberg Fachklinik Fasanenkiez Berlin; Berlin; Germany

40	Department of Women's and Children's Health; University of Padova; Padova; Italy

41	Department of Neuroscience; University of Padova; Padova; Italy

42	Department of Clinical Neuroscience; Karolinska Institutet; Stockholm; Sweden

43	Altrecht Eating Disorders Rintveld; Altrecht Mental Health Institute; Zeist; Utrecht; The Netherlands

44	Department of Child and Adolescent Psychiatry, Psychosomatics and Psychotherapy; LVR University Clinic Essen, University of Duisburg-Essen; Essen; Northrhine-Westfalia; Germany

45	College of Nursing; Seattle University; Seattle; Washington; United States

46	Nantes Université; CNRS, INSERM, l'institut du thorax; F-44000 Nantes; France

47	Department of Psychiatric Genetics, Medical Biology Center; Poznan University of Medical Sciences; Poznan; Poland

48	Department of Psychiatry and Behavioral Sciences; Stanford University; Stanford; California; United States

49	Department of Child and Adolescent Psychiatry, Psychosomatics and Psychotherapy, Center of Mental Health; University Hospital Wuerzburg; Würzburg; Bavaria; Germany

50	Department of Psychiatry; Reinier van Arkel; s-Hertogenbosch; Northern Brabant; The Netherlands

51	Department of Cancer, Epidemiology and Genetics; Masaryk Memorial Cancer Institute; Brno; Czech Republic

52	Department of Psychiatry and Psychotherapy, Comprehensive Center for Clinical Neurosciences and Mental Health (C3NMH); Medical University of Vienna; Vienna; Austria

53	First Department of Psychiatry; National and Kappodistrian University of Athens (NKUA); Athens; Greece

54	Department of Genetics; Queensland Institute of Medical Research QIMR Berghofer Medical Research Institute; Brisbane; Queensland; Australia

55	The Lundbeck Foundation Initiative for Integrative Psychiatric Research (iPSYCH); Aarhus University; Aarhus; Denmark

56	Center for Genomics and Personalized Medicine; Aarhus University; Aarhus; Denmark

57	Bioinformatics Research Centre; Aarhus University; Aarhus; Denmark

58	Department of Emergency and Post-Emergency Psychiatry; CHU, University of Montpellier; Montpellier; France

59	Helmholtz Zentrum München - German Research Centre for Environmental Health; Institute of Translational Genomics; Neuherberg; Germany

60	Human Genomics Research Group, Department of Biomedicine; University of Basel; Basel; Basel-Stadt; Switzerland

61	Department of Genomics, Life & Brain Center; University of Bonn; Bonn; Northrhine-Westfalia; Germany

62	Institute of Human Genetics; University of Bonn, School of Medicine & University Hospital Bonn; Bonn; Northrhine-Westfalia; Germany

63	Eating Disorders Unit; Parkland-Klinik; Bad Wildungen; Germany

64	Department of Clinical Psychology; University Hospital Bellvitge; Hospitalet del Llobregat (Barcelona); Barcelona; Catalonia; Spain

65	Department of Clinical Sciences; School of Medicine and Health Sciences; University of Barcelona; Hospitalet del Llobregat (Barcelona); Barcelona; Catalonia; Spain

66	Ciber Physiopathology of Obesity and Nutrition (CIBERObn); Instituto de Salud Carlos III; Madrid; Spain

67	Psychoneurobiology of Eating and Addictive Behaviors Research Group; Bellvitge Biomedical Research Institute (IDIBELL); Hospitalet del Llobregat (Barcelona); Barcelona; Catalonia; Spain

68	Centre for Psychological Services; University of Barcelona; Barcelona; Catalonia; Spain

69	Rheumatology Research Group; Vall d'Hebron Research Institute; Barcelona; Catalonia; Spain

70	Department of Psychiatry; First Faculty of Medicine; Charles University and General University Hospital; Prague; Czech Republic

71	Institute of Public Health and Clinical Nutrition; University of Eastern Finland; Kuopio; Finland

72	Institute of Applied Health Sciences; University of Aberdeen; Aberdeen; Scotland; United Kingdom

73	Division of Psychological and Social Medicine and Developmental Neurosciences; Faculty of Medicine; Technische Universität Dresden; Dresden; Germany

74	Department of Psychological Medicine; Stress, Psychiatry and Immunology Laboratory; Institute of Psychiatry, Psychology and Neuroscience; King's College London; London; United Kingdom

75	Center for Applied Genomics; Children's Hospital of Philadelphia; Philadelphia; Pennsylvania; United States

76	Division of Human Genetics; Children's Hospital of Philadelphia; Philadelphia; Pennsylvania; United States

77	Department of Pediatrics; University of Pennsylvania Perelman School of Medicine; Philadelphia; Pennsylvania; United States

78	Clinical Psychology Program; The Chicago School, Washington DC, College of Clinical Psychology; Washington DC; United States

79	Department of Psychiatry; University of Campania "Luigi Vanvitelli"; Naples; Italy

80	Department of Medical Research; Vestre Viken Hospital Trust, Bærum Hospital; Gjettum; Norway

81	Division of Mental Health and Addiction; NORMENT KG Jebsen Centre; Oslo University Hospital; Oslo; Norway

82	Padova Neuroscience Center; University of Padova; Padova; Italy

83	Department of Pathology and Biomedical Science; University of Otago; Christchurch; New Zealand

84	National Center for PTSD; VA Boston Healthcare System; Boston; Massachusetts; United States

85	Department of Psychiatry; Boston University Chobanian & Avedisian School of Medicine; Boston; Massachusetts; United States

86	Department of Mental and Physical Health and Preventive Medicine; University of Campania "Luigi Vanvitelli"; Naples; Italy

87	Department of Population Health; Queensland Institute of Medical Research QIMR Berghofer Medical Research Institute; Brisbane; Queensland; Australia

88	Department of Medicine Solna; Division of Rheumatology; Karolinska Institutet; Stockholm; Sweden

89	School of Nursing; University of North Carolina at Chapel Hill; Chapel Hill; North Carolina

90	Department of Psychiatry; Division of Psychiatric Genomics; Icahn School of Medicine at Mount Sinai; New York; New York; United States

91	Department of Genetics and Genomic Sciences; Mindich Child Health & Development Institute; Friedman Brain Institute; Icahn School of Medicine at Mount Sinai; New York; New York; United States

92	Department of Psychiatry; Helsinki University Hospital; Helsinki; Finland

93	Department of Public Health; University of Helsinki; Helsinki; Finland

94	Department of General Practice & Primary Healthcare, Faculty of Medical & Health Sciences; The University of Auckland; Auckland; New Zealand

95	Centre for Research in Eating and Weight Disorders, Department of Psychological Medicine; Institute of Psychiatry, Psychology and Neuroscience; King's College London; London; United Kingdom

96	Faculty of Medicine; Sigmund Freud University; Vienna; Austria

97	Institute of Biology and Medical Genetics; First Faculty of Medicine; Charles University; Prague; Czech Republic

98	Department of Child and Adolescent Psychiatry; Poznan University of Medical Sciences; Poznan; Poland

99	Department of Neuroscience, Psychology, Drug Research and Child Health (NEUROFARBA); University of Florence; Florence; Italy

100	Department of Psychiatry; University of Perugia; Perugia; Italy

101	Adolescent Health Unit, Second Department of Pediatrics, "P. & A. Kyriakou" Children's Hospital; National and Kappodistrian University of Athens (NKUA); Athens; Greece

102	Department of Clinical Psychology, Faculty for Social Sciences; Utrecht University; Utrecht; Utrecht; The Netherlands

103	Eating Disorders Unit, Department of Child and Adolescent Psychiatry; Medical University of Vienna; Vienna; Austria

104	Discipline of Psychology; Curtin University; Perth; Western Australia; Australia

105	Department of Translational Neuroscience; UMC Utrecht Brain Center; University Medical Center Utrecht, Utrecht University; Utrecht; Utrecht; The Netherlands

106	Department of Physiology; Institute of Neuroscience and Physiology; Sahlgrenska Academy at University of Gothenburg; Gothenburg; Sweden

107	Institute of Environmental Medicine; Karolinska Institutet; Stockholm; Sweden

108	Centre for Occupational and Environmental Medicine; Region Stockholm; Stockholm; Sweden

109	Centre for Precision Psychiatry; University of Oslo; Oslo; Norway

110	KG Jebsen Centre for Neurodevelopmental Disorders Research; University of Oslo; Oslo; Norway

111	Eating Recovery Center; Hunt Valley; Maryland; United States

112	Department of Psychiatry; ERC Pathlight; University of Maryland, St. Joseph Medical Center; Baltimore; Maryland; United States

113	Department of Psychiatry; University of Minnesota; Minneapolis; Minnesota; United States

114	The Weindrich Department of AI and Human Health; Icahn School of Medicine at Mount Sinai; New York; New York; United States

115	Department of Psychiatry; Icahn School of Medicine at Mount Sinai; New York; New York; United States

116	Department of Psychosomatic Medicine and Psychotherapy; Hannover Medical School; Hannover; Germany

117	Department of Nutrition and Dietetics; Harokopio University; Athens; Greece

118	Department of Psychiatry; Rutgers University; Piscataway; New Jersey; United States

119	Eating Disorder Research and Treatment Center, Department of Child and Adolescent Psychiatry; Faculty of Medicine; Technische Universität Dresden; Dresden; Germany

120	Research Department; Quantitative Genomics Laboratories (qGenomics); Barcelona; Catalonia; Spain

121	Institute of Mathematics and Statistics; University of Tartu; Tartu; Estonia

122	Institute of Neuroscience and Medicine (INM-1); Research Center Juelich; Juelich; Germany

123	Centre for Human Genetics; University of Marburg; Marburg; Germany

124	Université Paris Cité, INSERM U1266 (IPNP); Institute of Psychiatry and Neuroscience of Paris; Paris; Ile de France; France

125	Sainte-Anne hospital (CMME); GHU Paris Psychiatrie et Neurosciences; Paris; Ile de France; France

126	Department of Child and Adolescent Psychiatry, Psychosomatics and Psychotherapy; RWTH Aachen University; Aachen; Germany

127	Biological Psychiatry Laboratory; McLean Hospital; Harvard Medical School; Belmont; Massachusetts; United States

128	Eating Recovery Center; Denver; Colorado; United States

129	Department of Psychological Medicine; University of Otago; Christchurch; New Zealand

130	Specialist Mental Health Clinical Research Unit; Health New Zealand - Canterbury; Christchurch; New Zealand

131	Department of Psychiatry; Centre for Addiction and Mental Health; University of Toronto; Toronto; Ontario; Canada

132	Department of C & A Psychiatry; Medical University of Vienna; Vienna; Austria

133	Groningen Institute for Evolutionary Life Sciences; University of Groningen; Groningen; The Netherlands

134	Department of Psychiatry; University of California San Diego; San Diego; California; United States

135	Department of Psychiatry; University of Toronto; Toronto; Ontario; Canada

136	Tanenbaum Centre; Centre for Addiction and Mental Health; Toronto; Ontario; Canada

137	Department of Psychiatry; Ilsan Paik Hospital, Inje University; Goyang; South Korea

138	Department of Psychology; Michigan State University; East Lansing; Michigan; United States

139	Department of Psychiatry and Neurochemistry; Institute of Neuroscience and Physiology; University of Gothenburg; Gothenburg; Sweden

140	Department of Clinical Science; University of Bergen; Bergen; Norway

141	Maria Sklodowska-Curie National research Institute of Oncology; Warsaw; Poland

142	InsideOut Institute; University of Sydney; Sydney; Australia

143	Department of Community Health and Epidemiology; Dalhousie University; Halifax; Nova Scotia; Canada

144	Institute of Psychiatric Phenomics and Genomics (IPPG); Ludwig-Maximilians-Universität München; Munich; Germany

145	Department of Mental Health and Neuroscience; Queensland Institute of Medical Research QIMR Berghofer Medical Research Institute; Brisbane; Queensland; Australia

146	School of Psychology; University of Queensland; Brisbane; Queensland; Australia

147	School of Psychology and Counselling; Queensland University of Technology; Brisbane; Queensland; Australia

148	Great Ormond Street Institute of Child Health; University College London; London; United Kingdom

149	Department of Psychiatry and Behavioral Science; University of North Dakota; Fargo; North Dakota; United States

150	Department of Medicine, Surgery and Dentistry "Scuola Medica Salernitana"; University of Salerno; Salerno; Italy

151	Department of Psychiatry and Biobehavioral Sciences; University of California Los Angeles; Los Angeles; California; United States

152	Université Paris Cité; Paris; Ile de France; France

153	INSERM U1266; INSERM U1266; Paris; Ile de France; France

154	Institute of Clinical Medicine; University of Oslo; Oslo; Norway

155	Department of Health Sciences; University of Florence; Florence; Italy

156	German Center for Mental Health (DZPG); Berlin-Potsdam; Germany

157	Department of Psychiatry and Psychotherapy; Charité - Universitätsmedizin; Berlin; Germany

158	Department of Adult Psychiatry; Poznan University of Medical Sciences; Poznan; Poland

159	The Centre for Applied Genomics, Program in Genetics and Genomic Biology; The Hospital for Sick Children; Toronto; Ontario; Canada

160	McLaughlin Centre and Department of Molecular Genetics; University of Toronto; Toronto; Ontario; Canada

161	GGZ Rivierduinen Eating Disorders Ursula; Leiden; The Netherlands

162	Department of Psychiatry; Leiden University Medical Centre; Leiden; The Netherlands

163	Department of Environmental Epidemiology; Nofer Institute of Occupational Medicine; Lodz; Poland

164	Discipline of Psychology; Flinders Institute for Mental Health and Wellbeing; Adelaide; South Australia; Australia

165	Department of Clinical Medicine; University of Copenhagen; Copenhagen; Denmark

166	Department of Psychosomatic Medicine and Psychotherapy; University Medical Hospital Tuebingen; Tuebingen; Germany

167	German Centre for Mental Health, Tuebingen; University Tuebingen; Tuebingen; Germany

168	Department of Nutrition; University of North Carolina at Chapel Hill; Chapel Hill; North Carolina; United States

169	Psychiatry Department; McGill University; Montreal; Quebec; Canada

170	Eating Disorders Continuum; Douglas Mental Health University Institute; Montreal; Quebec; Canada

171	Department of Neuroscience; Janssen Research & Development, LLC; Titusville; New Jersey; United States

172	Department of Psychiatry; Amsterdam University Medical Center; Amsterdam; The Netherlands

173	Department of Psychiatry and Neuropsychology; School for Mental Health and Neuroscience, Maastricht University Medical Center; Maastricht; The Netherlands

174	GGZ InGeest; Amsterdam; The Netherlands

175	Professor of Medicine; University of Colorado School of Medicine; Aurora; Colorado; United States
